# Use of the Elimination Strategy in Response to the COVID-19 Pandemic: Health and Economic Impacts for New Zealand Relative to Other OECD Countries

**DOI:** 10.1101/2021.06.25.21259556

**Authors:** Nick Wilson, Leah Grout, Jennifer A Summers, Nhung Nghiem, Michael G Baker

## Abstract

**Background:** In response to the COVID-19 pandemic, some countries in the Asia-Pacific Region used very intensive control measures, and one of these, New Zealand (NZ), adopted a clear “elimination strategy”. We therefore aimed to compare key health and economic outcomes of NZ relative to OECD countries as of mid-June 2021.

**Methods:** This analysis compared health outcomes (cumulative death rates from COVID-19 and “excess death” rates) and economic measures (quarterly GDP and unemployment levels) across OECD countries.

**Results:** NZ had the lowest cumulative COVID-19 death rate in the OECD at 242 times lower than the 38-OECD-country average: 5·2 vs 1256 per million population. When considering “excess deaths”, NZ had the largest negative value in the OECD, equivalent to around 2000 fewer deaths than expected. When considering the average GDP change over the five quarters of 2020 to 2021-Q1, NZ was the sixth best performer (at 0·5% vs -0·3% for the OECD average). The increase in unemployment in NZ was also less than the OECD average (1·1 percentage points to a peak of 5·2%, vs 3·3 points to 8·6%, respectively).

**Conclusions:** New Zealand’s elimination strategy response to COVID-19 produced the best mortality protection outcomes in the OECD. In economic terms it also performed better than the OECD average in terms of adverse impacts on GDP and employment. Nevertheless, a fuller accounting of the benefits and costs needs to be done once the population is vaccinated and longer-term health and economic outcomes are considered.

## INTRODUCTION

As with a number of other jurisdictions in the East Asia and Pacific region, New Zealand adopted tight border controls and other stringent public health and social measures (PHSMs) in response to the COVID-19 pandemic.^1^ New Zealand also articulated an unambiguous COVID-19 elimination strategy.^2^ This strategy was successful,^3^ despite occasional border system failures that have caused outbreaks,^4^ following which its elimination status has been regained in each instance (up to at least mid-June 2021).

Other Organisation for Economic Cooperation and Development (OECD) countries have used suppression/mitigation strategies with poorer results (with these strategies defined elsewhere^5^). Nevertheless, the Australian strategy of “aggressive suppression” has also achieved COVID-19 elimination, albeit also with episodic outbreaks from border system failures.^4^ As of mid-June 2021, it also appeared that Iceland had eliminated community transmission of SARS-CoV-2.^6^

The COVID-19 pandemic has also generated major economic impacts globally and in OECD countries. For New Zealand, the economic impacts were driven by nearly a complete end to arrivals of international tourists and international students during most of 2020-21. There have also been labour shortages in various economic sectors (eg, of migrant workers for the agricultural and hospitality sectors). Countering the economic impact has been the New Zealand Government’s response with a very large fiscal package to subsidise wages and create new jobs. Additionally, a quarantine-free travel zone opened up in April 2021 with Australia and the Cook Islands, and the country may also have benefited from highly skilled citizens who returned to live in New Zealand.

Given this background, we aimed to compare key health and economic outcomes of New Zealand’s response to the COVID-19 pandemic using an elimination strategy, relative to OECD countries (as of mid-June 2021).

## METHODS

### Health outcomes

Health impact comparisons were made between New Zealand and other OECD countries/country groupings (with the OECD comprising 38 countries, which are largely high-income ones). The COVID-19 cumulative death rate data (including country populations) was for 11 June 2021 from the Worldometers website^7^ and was for probable and confirmed deaths.

Since analysis of “excess deaths” may provide a more accurate estimate of COVID-19 deaths,^8^ we also considered “excess deaths” as calculated by The Economist.^9^ These were for the period from when 50 COVID-19 deaths were reported in each country (typically March 2020), up to early 2021 (typically April 2021). The business-as-usual comparator for this analysis was based on a statistical model which predicted the number of deaths that might normally have been expected in each country. This model fitted a linear trend to years, to adjust for long-term increases or decreases in deaths, and a fixed effect for each week or month. The model was based on 121 indicators for over 200 countries and territories and involved a machine-learning model using gradient boosting. A description of the methods, the data and replication code are all available online.^10^

To estimate the number of life-years saved by the New Zealand response, relative to the OECD average, we considered two estimates from other OECD countries that we identified in the literature. One was for England which estimated the average quality-adjusted life years (QALY) lost per person dying from COVID-19 at 5·9 years.^11^ The other was from a German study which estimated an average of 9·6 years of life lost with a premature death from COVID-19.^12^

### Economic outcomes

Data on quarterly gross domestic product (GDP) changes, using percentage change relative to the previous quarter, were obtained from the OECD for 2020 and the first quarter of 2021.^13^ The only value missing from the latter period in the OECD dataset was the value for New Zealand and this was obtained from an official government website.^14^ Quarterly unemployment data was also obtained from the OECD.^15^ For New Zealand, *underemployment* data (officially described as “underutilisation”) were from official government statistics.^16^

## RESULTS

### Health outcomes

As of 11 June 2021, New Zealand had the lowest cumulative COVID-19 death rate in the OECD (5·2 per million population [26 deaths], Table 1). This rate was 242 times lower than the 38-OECD-country average (1256 per million). Australia had the second lowest COVID-19 death rate in the OECD, albeit seven times that of New Zealand (36 vs 5·2 per million). The best performing country grouping was the Asia-Pacific group of countries, followed by Nordic countries, and then island nations (Table 1, Figure 1).

**Table 1:**
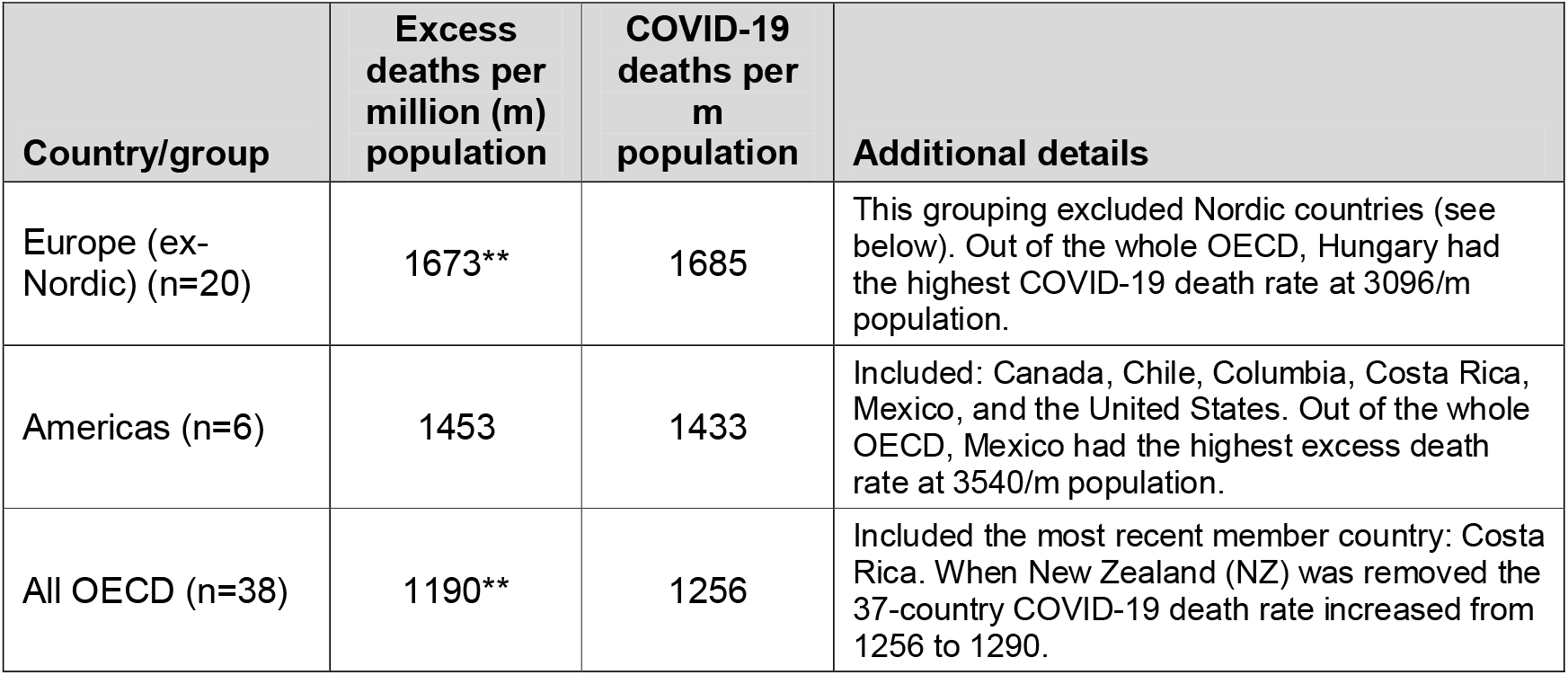
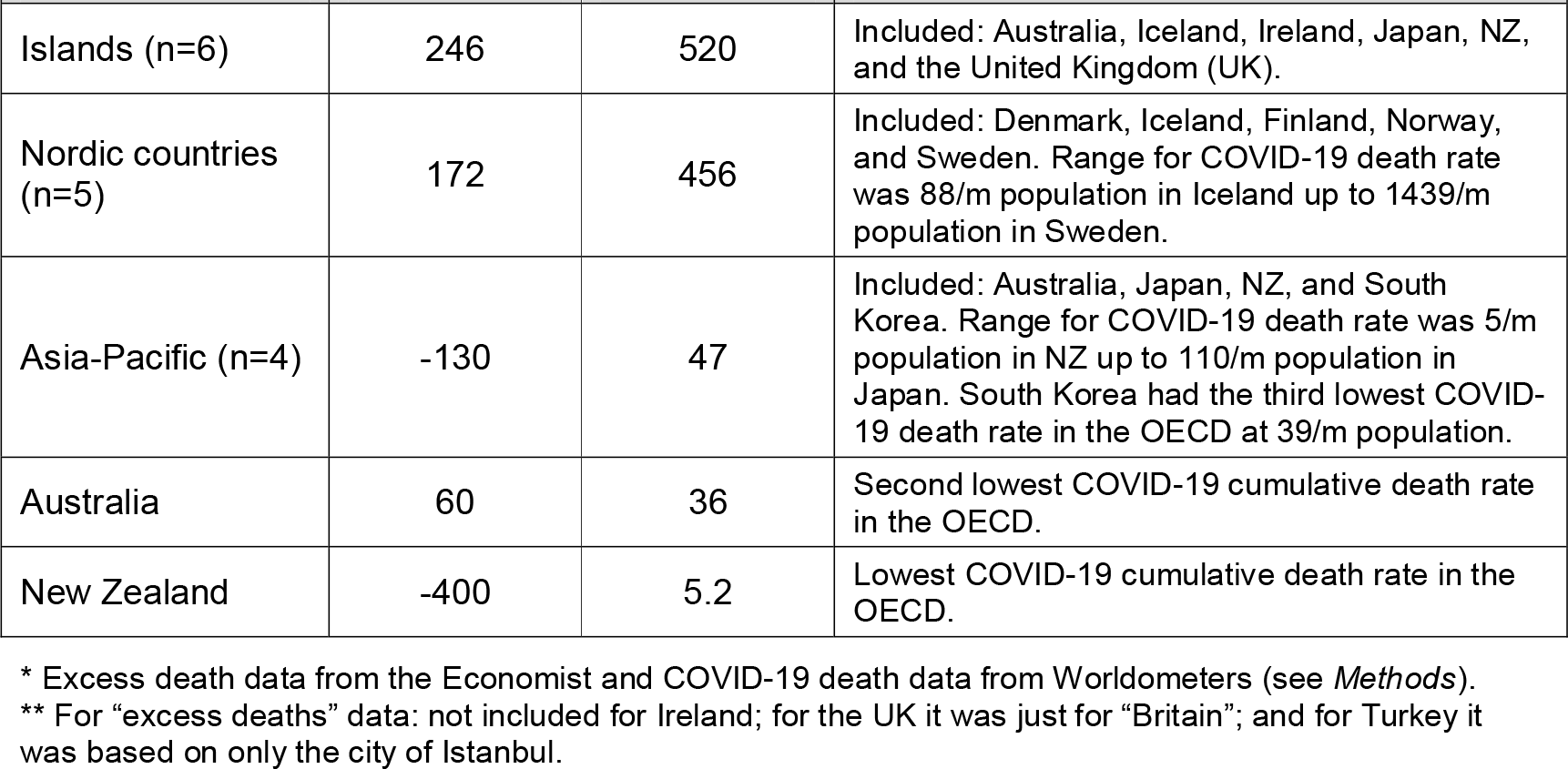
Average cumulative excess death rate and cumulative COVID-19 death rate in OECD countries and country groupings, as of June 2021*

| Country/group | Excess deaths per million (m) population | COVID-19 deaths per m population | Additional details |
| --- | --- | --- | --- |
| Europe (ex-Nordic) (n=20) | 1673** | 1685 | This grouping excluded Nordic countries (see below). Out of the whole OECD, Hungary had the highest COVID-19 death rate at 3096/m population. |
| Americas (n=6) | 1453 | 1433 | Included: Canada, Chile, Columbia, Costa Rica, Mexico, and the United States. Out of the whole OECD, Mexico had the highest excess death rate at 3540/m population. |
| All OECD (n=38) | 1190** | 1256 | Included the most recent member country: Costa Rica. When New Zealand (NZ) was removed the 37-country COVID-19 death rate increased from 1256 to 1290. |
| Islands (n=6) | 246 | 520 | Included: Australia, Iceland, Ireland, Japan, NZ, and the United Kingdom (UK). |
| Nordic countries (n=5) | 172 | 456 | Included: Denmark, Iceland, Finland, Norway, and Sweden. Range for COVID-19 death rate was 88/m population in Iceland up to 1439/m population in Sweden. |
| Asia-Pacific (n=4) | -130 | 47 | Included: Australia, Japan, NZ, and South Korea. Range for COVID-19 death rate was 5/m population in NZ up to 110/m population in Japan. South Korea had the third lowest COVID-19 death rate in the OECD at 39/m population. |
| Australia | 60 | 36 | Second lowest COVID-19 cumulative death rate in the OECD. |
| New Zealand | -400 | 5.2 | Lowest COVID-19 cumulative death rate in the OECD. |

\* Excess death data from the Economist and COVID-19 death data from Worldometers (see *Methods*).
\*\* For “excess deaths” data: not included for Ireland; for the UK it was just for “Britain”; and for Turkey it was based on only the city of Istanbul.

**Figure 1:**
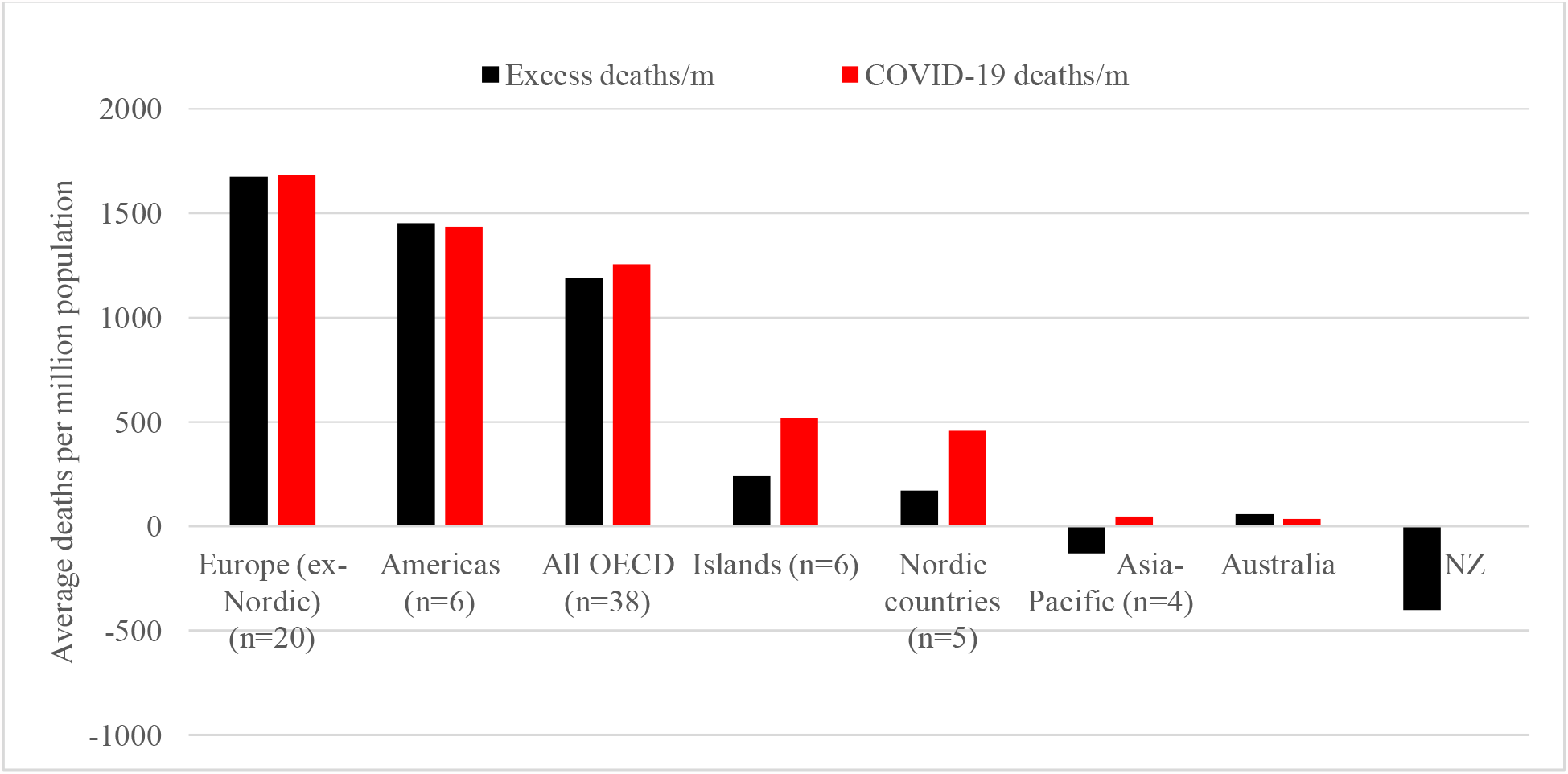
Average cumulative excess death rate and cumulative COVID-19 death rate in OECD countries and country groupings, as of June 2021 (see Table 1 and Methods for specific details)

From these results we estimate that, compared to the OECD average death rate from COVID-19, New Zealand avoided around 6430 deaths from COVID-19 as of mid-June 2021. Furthermore, based on QALY loss per premature death from COVID-19 (using data from England), this is equivalent to preventing a loss of 38,100 QALYs. But if data from Germany is used for this estimate, then a total of 61,700 life years were saved.

When considering “excess deaths”, New Zealand had the largest negative value in the OECD. That is, there was actually a decline in the death rate of 400 per million population (Table 1, Figure 1), equivalent to around 2000 fewer deaths compared to previous years. This pattern of a “negative excess death rate” was also reported for three Nordic countries (Denmark, Iceland, and Norway), and for two Asian countries (Japan and South Korea).

### Economic impacts (GDP)

New Zealand’s tight border restrictions and stringent lockdown in response to the initial outbreaks in March/April 2020 resulted in an overall economic downturn in the first half of the 2020 year (Figure 2). But there was a marked rebound in the third quarter (Q3) of 13·9% and positive growth at 1·6% in the most recent quarter for which data were available (2021-Q1). Overall the average GDP change over the five quarters studied meant that New Zealand was the sixth best performer in the OECD (at 0·5% vs -0·3% for the OECD average). Nevertheless, better performing countries for this same metric were: Ireland (1·7%), Turkey (1·7%), Chile (0·9%), Luxembourg (0·8%), and Estonia (0·7%). Australia also did fairly well at eighth in the OECD (0·2%). The poorest performing OECD country was Iceland (−2·1%). Nevertheless, the worst quarterly change for New Zealand was poorer than the OECD average (−11·0% vs -9·7%, respectively). However, the largest such quarterly change for an OECD country was for the UK (−19·5%).

**Figure 2:**
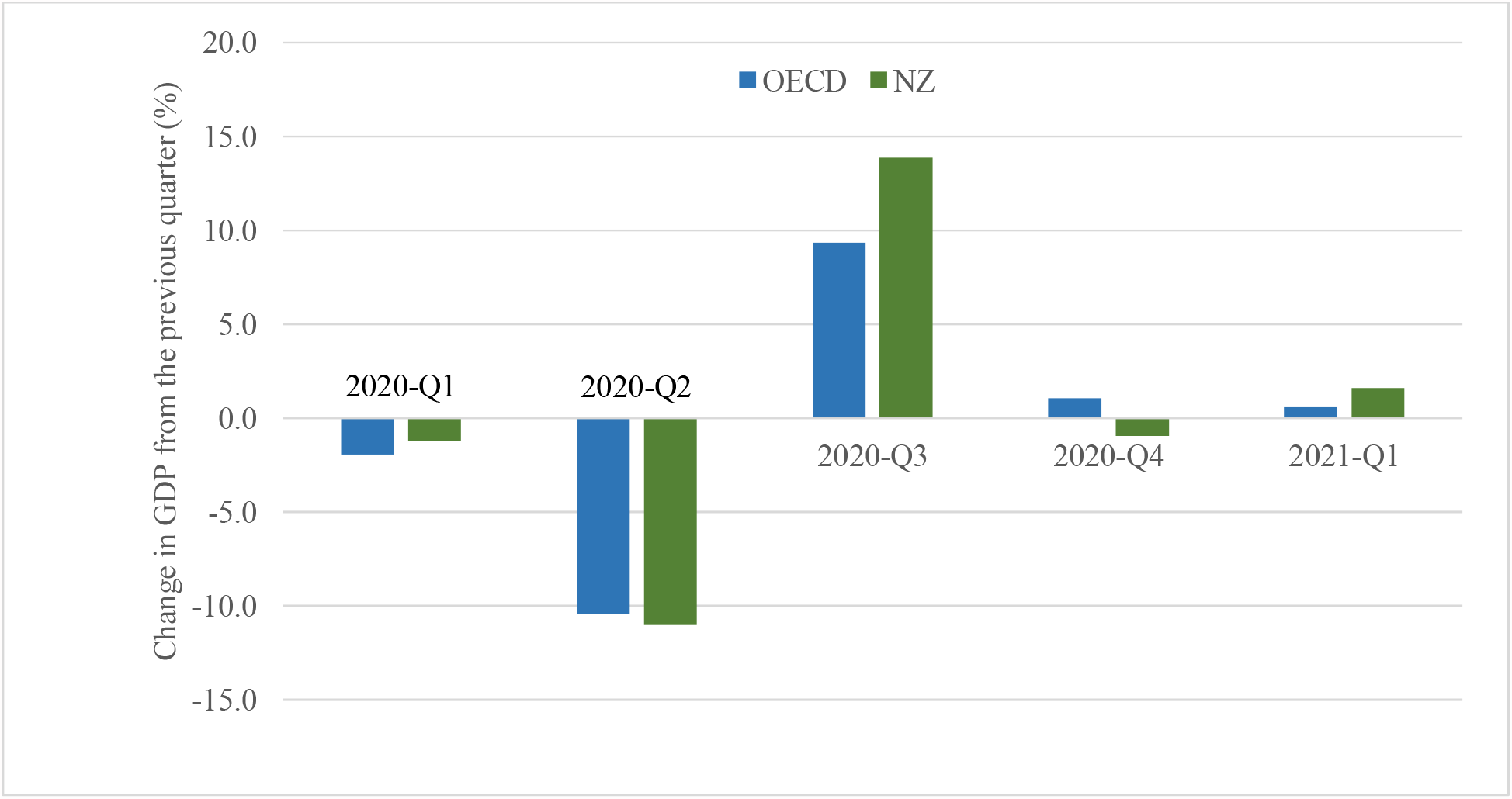
Changes in quarterly GDP for the OECD and New Zealand (see Methods for specific details)

### Economic impacts (employment)

In terms of unemployment, the New Zealand levels increased by 1·1 percentage points from before the pandemic (2019-Q4) to a peak of 5·2% in the third quarter of 2020 (Figure 3). This was less than the respective values for the OECD of 3·3 points to 8·6% (in 2020-Q2). In the last quarter of 2020, New Zealand had the tenth lowest unemployment level, and in the ninth lowest for the first quarter of 2021 (albeit data only available for 34 OECD countries for the latter).

**Figure 3:**
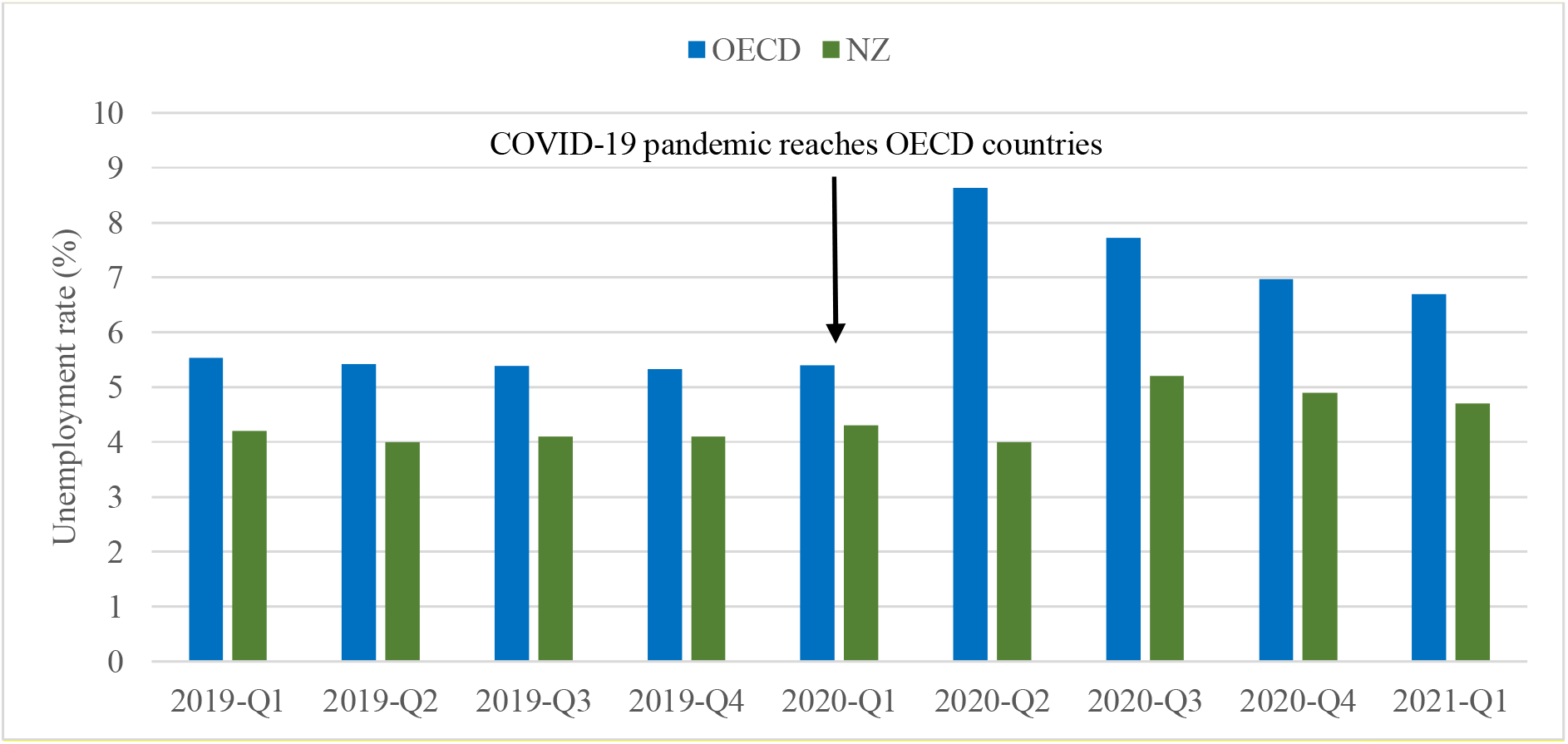
Unemployment rate (%) in the OECD and New Zealand.

Underemployment data for New Zealand showed an increased from 10·1% in the last quarter of 2019 up to a peak of 13·1% in the third quarter of 2020. For the first quarter of 2021 it had declined to 12·2% overall albeit with differences by sex (14·7% for women, 10·1% for men). Also it was underemployment in women that increased the most at 16.4% in the second quarter of 2020 (vs the highest increase of 12.9% increase for men in the third quarter of 2020) (Figure 4).

**Figure 4:**
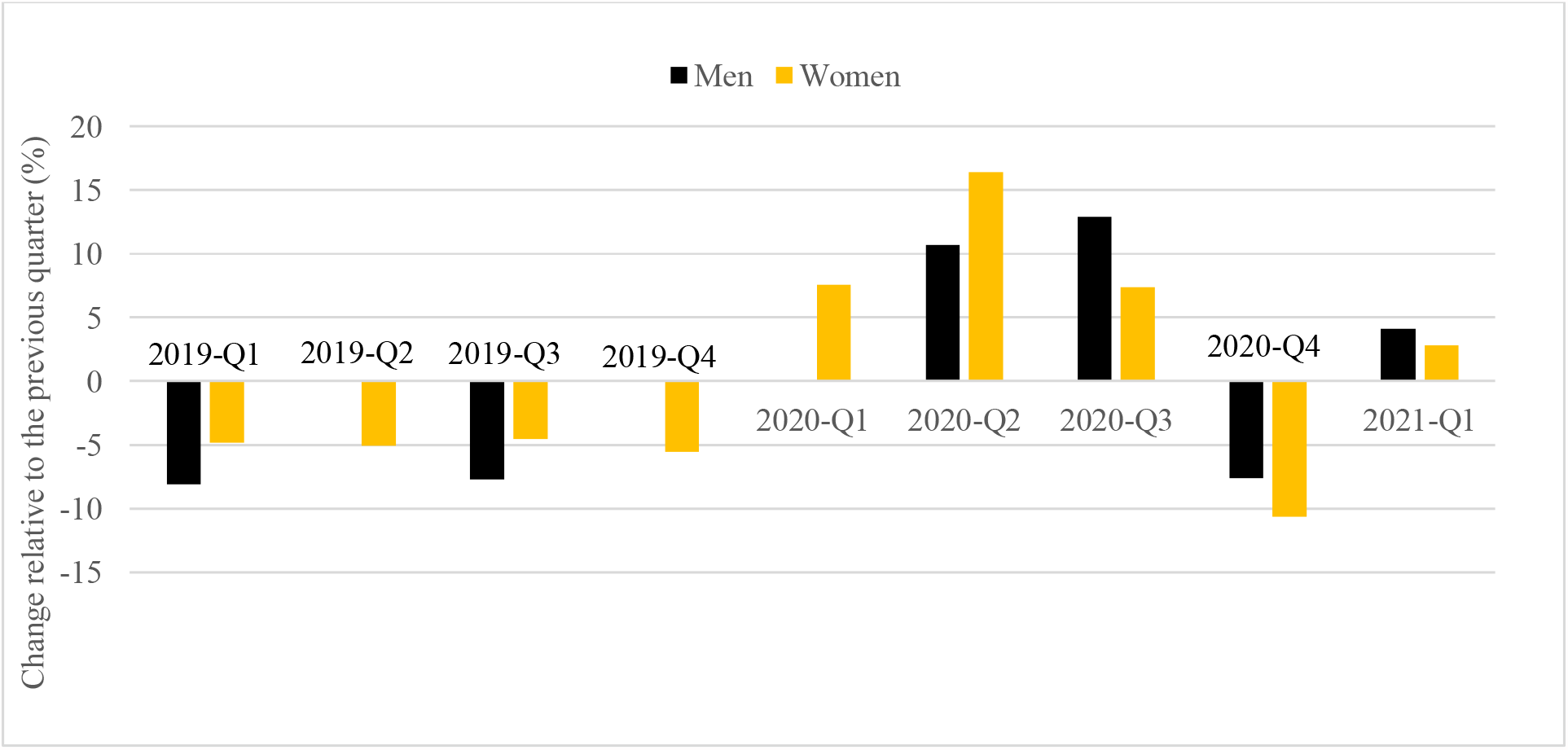
Changes in the underemployment rate (%) in New Zealand.

## DISCUSSION

### Main findings

These results indicate that New Zealand’s successful COVID-19 elimination strategy delivered more mortality protection benefits than any other OECD country. Indeed, the control measures reduced the death rate to below the typical pre-pandemic level – with this outcome probably due to reductions in deaths from seasonal influenza and other viral infections (ie, with international declines seen for respiratory illness from: influenza virus, parainfluenza virus, respiratory syncytial virus, metapneumovirus, and non-SARS-CoV-2 coronaviruses^17^; as well as in New Zealand^18^). Other researchers have reported that New Zealand had the lowest excess death rate in 2020 out of 29 high-income countries, and actually had an estimated 2500 fewer deaths than expected (95%CI: 2100 to 2900).^19^ Another study reported that New Zealand (along with: Denmark, Finland, Norway, South Korea, and Taiwan) were jurisdictions where life expectancy actually increased between 2018 and 2020, in contrast to many other high-income countries where decreases occurred because of the pandemic.^20^

Collectively these results provide support for the benefit of adopting an elimination strategy towards this pandemic, in preference to a suppression/mitigation strategy, as also concluded elsewhere.^5 21 22^ Nevertheless, such a pandemic response strategy may be more achievable for countries with the necessary competent leadership, as described for New Zealand,^23^ and in comparisons between the New Zealand and United States response.^24^ Furthermore, “conditions favouring successful elimination include informed input from scientists, political commitment, sufficient public health infrastructure, public engagement and trust, and a safety net to support vulnerable populations.”^5^ These factors may also have been relevant to the relative successes of Australia and Iceland, albeit with a particularly strong science-based response in Iceland.^25^

Being an island nation probably facilitated relatively good border control in New Zealand and two other relatively successful OECD countries: Australia and Iceland (Table 1). Similarly, for South Korea which can be considered a “virtual island” due to its militarised land border with North Korea. In contrast, however, the islands of Ireland and Britain did not appear to make good use of border controls at an early stage. Also of note is that the non-OECD countries of China and Vietnam succeeded during 2020 with apparent elimination strategies despite having very long land borders.

### Study strengths and limitations

This analysis benefited from the case study country (New Zealand) having a very clearly articulated elimination strategy, whereas the strategic approach is generally poorly defined for other OECD countries (ie, varying intensities of suppression and mitigation^5^ and sometimes shifting from mitigation to suppression as per the UK). The analysis was also able to supplement COVID-19 death rate data with “excess deaths” data to obtain a richer understanding of the likely health impacts. Focusing on OECD countries also allows for relatively standardised comparisons of economic data.

Nevertheless, this analysis is both preliminary and fairly high-level. The pandemic is far from over globally and New Zealand could yet experience border system failures that result in major health and economic consequences (eg, before a high proportion of the population is vaccinated). In particular, the further economic recovery for other OECD countries with higher vaccination levels than New Zealand, could change the longer-term economic analysis. That is, the ultimate economic assessment of the pandemic responses should probably consider a multi-year period.

More specific limitations that will have underestimated the health and economic benefits of New Zealand’s elimination strategy to date, include:

- Our analysis only considered life years or QALYs gained from the prevention of premature death from COVID-19. Yet data from the United States suggests that this is incomplete with an estimated 15% of QALY loss from COVID-19 being due to non-fatal illness and impact on families.^26^ Furthermore, this proportion may increase in the future once there is a better understanding of long-term sequelae (already described as “substantial health burden”^27^).
- We did not assess the morbidity reduction benefits of the non-COVID-19 health conditions prevented by the control measures (eg, the reduction of infectious respiratory diseases^18^ and injuries^28^ described in the New Zealand context).
- We did not quantify the potential benefits for protecting specific population groups from unequal harm, thereby exacerbating health inequities. For example, Māori (Indigenous population of New Zealand) have suffered excessive death rates in three previous pandemics.^29^

But on the other-hand, our analysis did not quantify additional downsides of New Zealand’s COVID-19 response:

- The harm to health from the disruption to routine health care provision, albeit the evidence to date seems to suggest minimal impact eg, for cancer care in New Zealand.^30^
- The psychological distress associated with lockdown responses required in March-May 2020.^31^ There was also an increase in alcohol-related emergencies involving ambulance staff attendances,^32^ and increased smoking levels in some groups.^33^
- However, some people reported that the lockdown experience had positive psycho-social aspects,^34^ and all these impacts need to be balanced with the psychological benefits from eliminating the risk of infection in the community. Also, the negative effects of the lockdown would have been much less in New Zealand, given the much shorter duration of lockdowns in this country,^1^ when compared with other OECD countries that pursued suppression/mitigation strategies.

We also used high-level economic metrics such as GDP which only partially relates to human wellbeing, and indeed the New Zealand Government has been shifting its focus from GDP to “wellbeing”. More specifically it uses a “Living Standards Framework”, which includes indicators across multiple domains, such as health, environment, cultural identity, social connections, and subjective wellbeing.^35 36^ Also the consideration of country-level GDP changes disguises particularly severe shocks to particular parts of the New Zealand economy (ie, the international tourism sector, the provision of education to international students, and sectors dependent on migrant workers). The GDP comparison between countries also did not allow for country size or trade dependency. For example, a country with a very large internal market such as the United States, is far less dependent on external trade, international tourism and migration than New Zealand is. On the other hand, New Zealand’s relative dependence on trade with China probably facilitated its economic recovery, given China’s early success in eliminating COVID-19 and its own rapid recovery. At least in 2021, the quarantine-free travel arrangements between Australia and New Zealand are also likely to be assisting economic recovery of both these nations.

Long-term economic benefits might also arise from New Zealand’s COVID-19 response. For example, more routine working from home might result in productivity gains for some sectors (and the benefits of reduced commuter traffic). A reduced dependence on low-cost foreign labour might also improve productivity in some sectors in the long-term, along with forcing the New Zealand Government to address “long-standing issues of disadvantage and under-delivery”.^37^ Also somewhat speculative is if New Zealand might ultimately benefit economically from some of its diaspora who have returned in a purported “brain gain” for the country.^38^

### Potential research and policy implications

A full and proper analysis of the “elimination strategy” response to the COVID-19 pandemic will need to take account of a much longer time period, and await a time until COVID-19 vaccination coverage has stabilised in the various OECD countries. In particular, New Zealand and Australia will need to identify an optimal long-term strategy for COVID-19. The broad options include accepting COVID-19 as an endemic infection, with ongoing vaccination to protect the most vulnerable (analogous to what is done for seasonal influenza). An alternative is sustained elimination using a mix of high vaccine coverage, combined with PHSMs to control outbreaks (much as is done for measles in OECD countries experiencing imported cases). Either way, these countries might need a period of carefully scaled-down border controls with only permitting quarantine-free travel to people who are vaccinated and who follow various processes before and after arrival (eg, regular tests, as modelled elsewhere^39^; use of digital tracking software; and avoidance of super-spreading settings). Assessing the risks and benefits of border control easing, including the risk of outbreaks and the economic harms of lockdowns, could benefit from integrated health and economic modelling analyses eg, as started for aspects of unemployment in New Zealand^40^ and for Victoria, Australia.^41^

There is even the possibility that the global community will adopt a COVID-19 goal of progressive elimination leading to global eradication,^42^ and then New Zealand and Australia could be part of expanding quarantine-free travel zones of other countries that have succeeded with elimination.^43^

## CONCLUSIONS

In terms of avoiding deaths from COVID-19 and overall reduction in excess deaths, New Zealand’s elimination strategy response to the pandemic was the best in the 38-country OECD grouping. In economic terms it also performed better than the OECD average in terms of impacts on GDP and employment. Nevertheless, a fuller accounting of the benefits and costs needs to be done once OECD country populations are vaccinated and longer-term health and economic outcomes are considered.

## Data Availability

The data used in the analysis is all available from public sources. Nevertheless, the relevant Excel spreadsheets are all available on request.

